# Person-Centred Critical Care: Lessons from a Service Evaluation

**DOI:** 10.1101/2024.01.26.24301616

**Authors:** S. Pearson, A. Petsas, J. Balabanovic, M. Juj, W. Harris, T. Bonnici

## Abstract

**Context:** ‘Critical care’ encompasses ‘intensive care’, ‘intensive therapy’ and ‘high dependency’ care and is operationalised when patients require specialised monitoring and intervention following complex surgery, or a life-threatening illness or injury.

**Background:** During the pandemic, the Critical Care Department at UCLH formed a family liaison team to bridge the connection gap between patients, families, and clinical teams. This evolved into the Patient & Family Team (PFT), which organised several engagement events to understand patient and family experiences in critical care.

**Methods:** Focus group discussions were conducted online and in-person with patients and bereaved families exploring their experience of the service. Discussions revolved around pivotal moments in the critical care journey. Feedback was analysed thematically and validated with the participants.

**Findings:** Patients described their journey through four stages: Admission, Period of Disorientation, Re-Awakening, and Recovery. Bereaved families categorised their experiences into seven stages from ‘The Phone Call’ to ‘Bereavement.’ The need for effective and compassionate communication and support was evident for both groups.

**Discussion:** Feedback revealed the emotional journeys of patients and families in critical care. While many experiences align with the existing literature, they also highlight areas for improvement, emphasising the value of human connection in healthcare. This study also demonstrated the need for continuous service evaluation and strategies for understanding underserved populations.

## Introduction

The COVID-19 pandemic exposed and exacerbated flaws and inequities that already existed in our healthcare system (Irizar et al. 2023, Anderson et al. 2023). We were exposed to the brutal reality of a healthcare system stripped of humanity and personhood.

Due to visiting limitations and the requirement to use personal protective equipment (PPE), patients and families were physically separated and cared for by faceless staff. Patients and families suffered. Staff also suffered, through moral injury and burnout due to the daily scale of distress witnessed and the inability to provide person-centred care due to workload pressures (Calkins et al. 2023). Many critical care staff have left the profession, replaced by younger and less experienced staff, some of whom only know nursing in the context of the pandemic (Vogt et al., 2023).

As we move into the post-pandemic era and observe the relaxation of pandemic-defined critical care restrictions, there is an urgent need to rebuild the connections between staff, patients, and families. It is essential to recognise that behind every role, be it patient, family member or staff, lies an individual with their own unique experiences and needs.

Person-centred care focuses on understanding and respecting everyone’s unique experiences, preferences, and values, fostering a collaborative and holistic approach to healthcare where all stakeholders, including caregivers and family members, are active participants in care decisions and planning (McCormack et al. 2010). By bridging the connection between patient satisfaction, job satisfaction, staff retention, and overall well-being, we can ensure better patient outcomes and enhance cost-effectiveness, efficiency, and the overall value provided to our healthcare system (Ulrich et al., 2014; Van Osch et al., 2017).

## Background

The University College London Hospitals NHS Foundation Trust (UCLH) is one of the largest NHS trusts in the United Kingdom with six critical care units (75 beds) providing specialist care including haematology, neurosurgery, urology, ENT, and maxillofacial care.

At the height of the pandemic, our critical care units were overwhelmed, like so many others. We were desperately striving to save lives and protect our staff. However, because of this urgency, the emotional and psychological care of patients, families, and even our staff was often overshadowed.

### The Family Liaison Team

In this challenging landscape, the Family Liaison Team (FLT) was formed in March 2020. Initially comprising of redeployed staff, this team became the bridge connecting patients, families, and the clinical team. As the crisis continued, the redeployed staff were replaced by volunteers. They facilitated video calls, visits, and provided that crucial touch of humanity. At its height, this team had up to seventy members, providing real-time public and patient engagement and adapting to feedback and changing circumstances on the fly.

Acknowledging the value of the insights gathered from the FLT and the pandemic’s experience, the UCLH Charity funded a dedicated team to assimilate these learnings into a sustainable and scalable model. With this, the evolution of the family liaison team extended its reach beyond supporting families alone, evolving into The Patient & Family Team (PFT) to encompass a holistic approach.

The Critical Care Patient & Family Team is the post-pandemic, business-as-usual re-incarnation of the Family Liaison team. The team is much smaller now, remains multidisciplinary, and continues to be supplemented by volunteers. We provide holistic and practical support for patients and families and work towards improving the overall patient experience and engagement.

## Aims and Objectives

The overall aim of the UCLH Charity funded project was, ‘To develop a model for person-centred care that delivers an excellent experience for patients, families and staff that is sustainable for the long term and is adaptable to local context.’

Anticipated quality outcomes by 2025 include:

- Improved patient, family, and staff experience, well-being, and mental health
- Reduction in complaints and conflicts among staff, patients and families
- Improved equity and inclusion of patients, families and staff
- Attraction of inward investment into the Trust and support for the innovation agenda
- Increased effectiveness of related trust-level initiatives in this domain

This paper reports the findings of the patient and family service evaluation engagement events that were facilitated during the project, conducted to gain a deeper understanding of patient and family experiences.

## Methods

The Patient & Family Team (PFT) embarked on a qualitative study to understand patient and family experiences in critical care. To achieve this, we invited patients discharged from critical care and bereaved families of patients who died in critical care within the past year to participate in engagement events. We intentionally excluded patients for whom participation might be inappropriate or distressing because of ongoing complaints or safeguarding concerns.

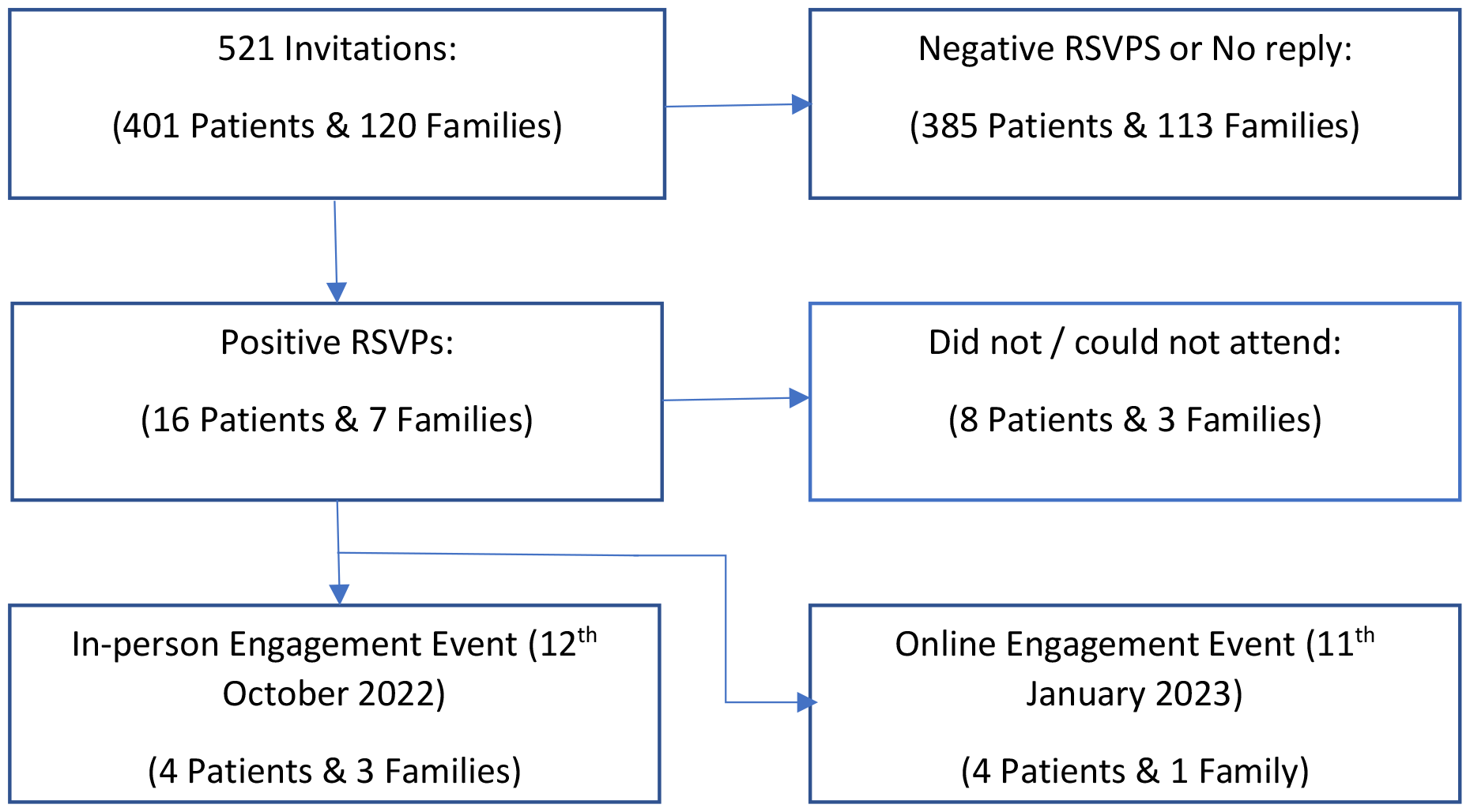

In line with guidance from the Health Research Authority (HRA) in England, this project was defined as service evaluation and not research (HRA, 2023), and therefore HRA approval was not required. However, to safeguard staff who participated in the evaluation it was conducted according to the United Kingdom Policy Framework for Health and Social Care Research (HRA, 2023), and approval to conduct the evaluation was given by local department governance and leadership. Information about the purpose of the engagement events was provided and participation was taken as implicit consent.

Two primary engagement events were scheduled, one in-person and one online via Zoom. Despite efforts to be inclusive, several patients and families could not participate due to timing or platform constraints. Some patients and families expressed interest in one-to-one interviews, however, lack of capacity meant that this could not be facilitated.

Each engagement session adopted a focus group format, built around a guided discussion of pivotal moments in the critical care journey (figure 1.). The groups were facilitated by a PFT member, with support from a Critical Care Psychologist, and another PFT member observed and documented the discussions. Patients (ICU survivors) and bereaved family members were separated into different focus groups.

**(Figure 1.).**
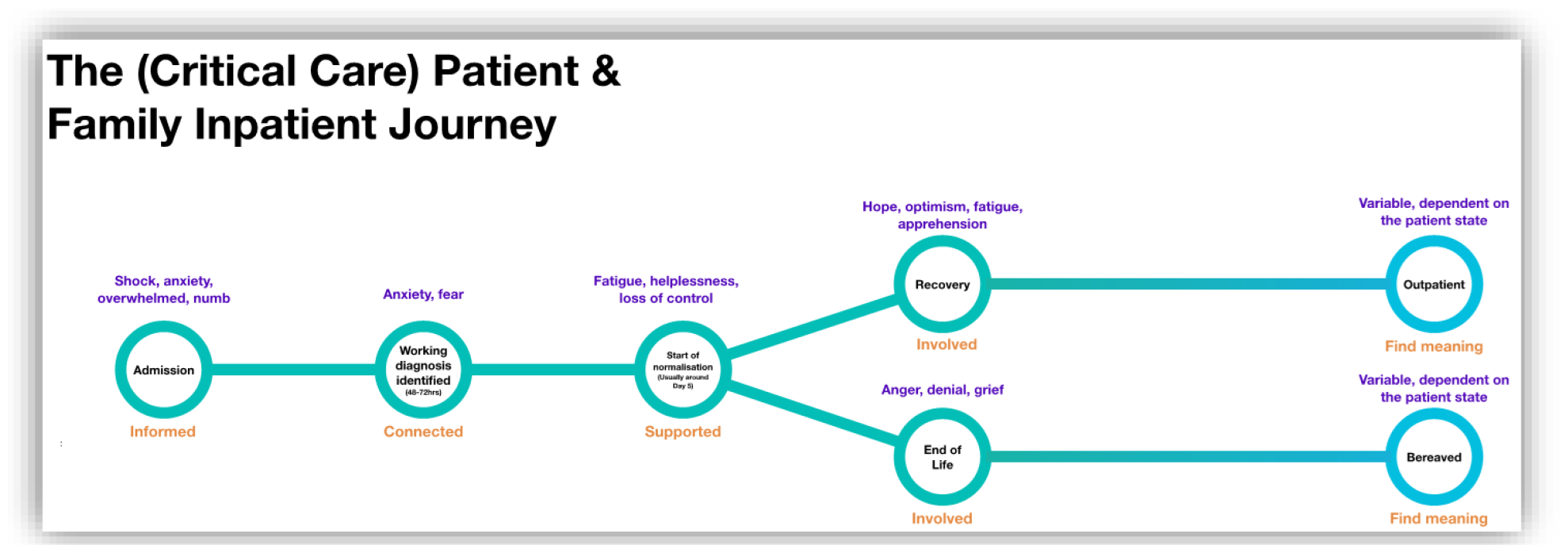

The events ran from 6 pm to 7:30 pm on a weekday, and the in-person event was held in the Education Centre at UCLH.

The objectives were to understand:

- The emotional and psychological experience of being a patient or relative/friend of a patient in critical care
- What went well for you while in critical care
- What the team could improve on

Following the event, the data were analysed collaboratively by members of the PFT and Psychology teams to minimize bias and to establish a consensus. To ensure the quality and relevance of our interpretations, we shared a summary of our findings with the participants for validation.

## Results

Two events were organized to gather insights. The first, an in-person gathering on 12th October 2022, was attended by four patients and three bereaved family members; the subsequent online session on 11th January 2023 saw participation from four patients, with one accompanied by a family member. To ensure a comprehensive understanding of the collected data, the findings have been separated into two primary categories: The Patient Journey and The Bereaved Family Journey, acknowledging the inherent differences and unique nuances of each experience.

### The Patient Journey

Despite the varied illnesses and treatments, we noticed a consistent narrative.

At the initial engagement event, the patients were presented with the patient and family inpatient journey (figure 1.) displaying what we believed to be the pivotal points. However, the diagram did not resonate with them, and through dialog, they constructed their journey through critical care in four interconnected stages (figure 2.).

**(Figure 2.).**
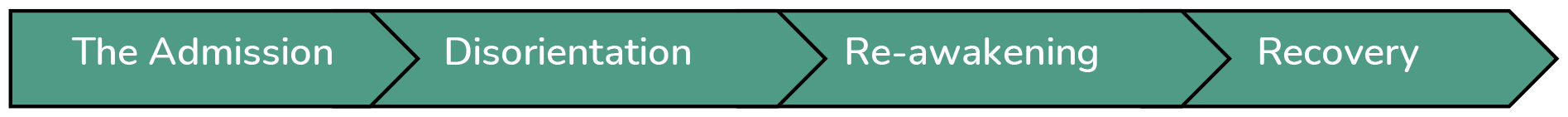

#### The Admission Phase

Patients who were aware of their admission experienced feelings of shock, fear, and anxiety. The need for information and reassurance was particularly important during this phase. One patient articulated upon their transfer to critical care, “*I don’t think there’s any way to eliminate the fear of going to ICU. But it’s good to know you’re in the right place, a safe place, and people are competent*.” Concerns were not confined to personal circumstances; many patients expressed anxiety about their families’ knowledge and well-being. To exemplify, a patient post-emergency surgery remarked, *“I didn’t know if my parents knew, I was most concerned that my parents didn’t know what had happened*.*”*

#### Period of Disorientation

The acute phase of critical illness was characterised by pronounced disorientation. Memories of delirium and hallucinations and feelings of confusion and fear were recurrent themes as described by one patient. *“I was so tired I had hallucinations and couldn’t sleep. I remember seeing someone sitting next to me, but I knew they were not really there*.*”* While some patients could rationalise these experiences, others found it challenging. The overarching sentiment was the need for reassurance, consistent support, and transparent communication.

#### Re-Awakening

Patients described a phase of ‘re-awakening’, a gradual return to self-awareness. The journey of being ‘pulled back’ into reality by staff, predominantly nurses, was highlighted *“The staff brought me back from sedation; they were my connection to the real world*.*”* Another patient observed, *“I remember feeling more “in the room” as I began to recover*.*”* Anecdotes from the outside were particularly appreciated, highlighting the importance of human connection. *“Hearing music and my nurse sharing stories from their life reminded me of the world outside, the life I would return to*.*”*

#### Recovery

Progressing toward recovery, patients highlighted feelings of boredom, monotony, and frustration due to lack of routine. During this phase, patients leaned heavily on staff interactions to maintain motivation. A patient reflected on this feeling of isolation, *“After ICU, I was moved to a side room on the ward. I felt forgotten, I stood at the door to watch people outside*.*”* The monotony of recovery was further emphasised by another patient who described their experience, *“After discharge from ICU, I went to the ward. It was mind-numbingly boring on the ward, very mundane. Nothing much happened from one day to the next*.*”* However, the pivotal role of nursing staff persisted as another patient recounted, “*The small tasks that the nurses would set me really helped. I wanted to please them by completing them*.”

#### Common Areas of Concern

Feedback also highlighted consistent negative experiences. Notably, these included feelings of vulnerability during shift transitions, perceived diminished staff availability during nights and weekends, recurrent sleep disturbances, and unanimous disapproval of the meals provided.

### The Bereaved Family Journey

Similar to the patients during the engagement event, the families felt that their experiences did not align with the journey, as shown in Figure 1. Their narratives can be broadly categorised into seven phases (figure 3.).

**figure 3.).**
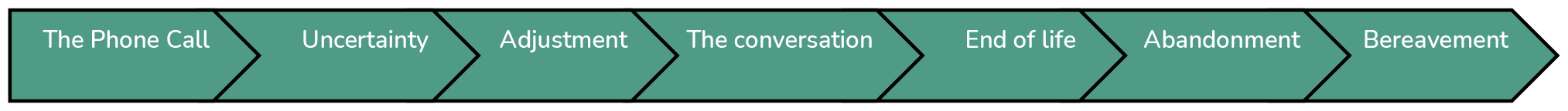

#### The Phone Cal

This stage is characterised by the sudden unexpected notification that a family member has been admitted to critical care. For many, this initial contact is disorienting, setting the stage for the difficult journey ahead.

#### Period of Uncertainty

During this phase, families struggle with an absence of comprehensive information, resulting in heightened feelings of vulnerability and helplessness. The situation is often highly anxiety-inducing, with many families searching for information to reduce the uncomfortable uncertainty.

#### Period of Adjustment

As time elapses, a pattern emerges. Families adjust to a new routine of hospital visits and medical updates. This is a time filled with a mixture of hope and helplessness, clinging to any signs of improvement or stability. Whereas patients talked about the bedside nurse providing the key therapeutic relationship, the key relationship with the bereaved families was formed with the doctors.

#### The Conversation

The communication of the inevitable, that their loved one will not survive, and comfort will be prioritised over treatment. It was described as a conversation that could never be unheard.

#### End of Life

This is the emotional stage in which families form a vigil around the patient and say their last goodbyes. This is a time of deep sorrow, potential regrets, and profound expressions of love.

#### Abandonment

There is an acute sense of isolation following the actual loss. Families often confront an overwhelming feeling of being ‘*dumped and abandoned*.*’* One participant poignantly recounted their solitary journey home from the hospital via the tube, leading to an empty house and solitude.

#### Bereavement

The concluding stage represents the formal recognition of loss. Rituals, ceremonies, and memorial services frequently accompany this phase, serving as both tribute to the departed and solace for the living.

## Discussion

The nature of critical illness, medical interventions, and the environment of critical care are inherently traumatic and dehumanising (Timmins et al., 2015). Fear and anxiety are common for both patients and families (Fumis et al., 2015; Freeman et al., 2021), but this study determined that family members of patients who died in critical care were more traumatized and vulnerable. This is consistent with studies that revealed higher levels of anxiety, depression, and post-traumatic stress symptoms among family members, particularly bereaved family members, than among patients (Fumis et al., 2015).

Both patients and bereaved families were uncomfortable with the critical care journey we presented to them for two reasons (figure 1.). First, the salient points from the service providers’ perspective did not resonate with their lived experiences, and second, the journey presented them with an alternate reality of either survival or death. The four distinct phases elucidated by the patients and the seven phases detailed by the bereaved families underscore the intricate, multi-layered experiences that individuals within both cohorts undergo.

For patients, the admission phase was a time of heightened anxiety and uncertainty. These findings are consistent with recent studies that highlight the profound psychological impacts of admission to critical care, including feelings of fear and anxiety (Vogel at al., 2023; Gil-Julia et al., 2020). Interestingly, on admission to critical care or when awaking from sedation, patients consistently voiced that their main concern was for their families, reiterating the interconnected emotional experiences of patients and their loved ones.

The bereaved family journey (figure 3.), highlights the importance of effective communication and support, outcomes consistent with previous studies (Turner-Cobb et al., 2016; Wong et al., 2015). Rarely is one prepared for the news that a loved one has been admitted to critical care (Gil-Julia et al., 2018). Stages such as ‘The Phone Call’ and ‘Period of Uncertainty’ draw attention to the acute need for timely, clear, and compassionate communication and information (Digby et al., 2023). The sentiments expressed during ‘The Conversation’ and ‘Abandonment’ phases highlight the need for sustained support, even after the death of the patient, which appears to be a crucial gap in many healthcare settings (Wearing, 2001). This is consistent with previous studies (Wong et al., 2015; Gil-Julia et al., 2018), which highlight the lasting trauma faced by families, especially in the absence of adequate psychological support.

The key relationship families formed with staff were with the doctors, which is consistent with other studies (O’Gara et al., 2021). This is understandable as doctors are often the key communicators at salient points in informing families of a loved one’s admission to critical care or communicating the futility of treatment (O’Gara et al., 2021). Patients, on the other hand, identified nurses as the key therapeutic relationship in their recovery from critical illness mirroring the Kuyler & Johnson (2023) study.

While critical care patients’ experiences of disorientation, confusion, hallucinations, and inadequate and interrupted sleep are well-documented (Koçak & Arslan, 2022; Gil-Julia et al., 2020), the emphasis on human connection in the ‘Re-awakening’ phase offers profound insights into the relationship between the patient and nurse. The anecdotes shared situate the nurse as more than a care provider, suggesting that they were pivotal in helping the patient to rediscover a sense of self and to re-establish personhood (Kuyler & Johnson, 2023).

In conclusion, our findings illuminate the nuanced, emotional, interconnected yet separate journeys undertaken by patients and their families during their critical care experiences (Flumis et al., 2015). While many experiences we heard resonated with established literature, they underlined the importance of human connection in healthcare and highlighted areas for service improvement.

## Limitations

The study acknowledges certain inherent limitations in its sample population. Notably, the participants in these events were predominantly white, well-educated, and proficient in English. This demographic skew may have influenced the nature of the feedback and experiences shared during the evaluation process. Consequently, the findings might not comprehensively represent the diverse range of experiences and challenges encountered by our patient population, particularly those from varied ethnic backgrounds, different educational levels, or with varying levels of language proficiency.

Furthermore, the narrative related to the bereaved families was primarily constructed through staff debriefings and reflections post-event, leading to an absence of direct quotations in the report. This contrasts with the patient group, where a facilitator transcribed the conversation. This methodological approach may limit the depth and authenticity of insights gathered from the bereaved families, potentially affecting the comprehensiveness of the study’s findings

## Conclusion

This service evaluation was conducted to gain a deeper understanding of the patient and family experience of critical care at UCLH. The feedback reveals salient points and stages of the critical care journey that are defined by and meaningful to patients and bereaved families. This provides invaluable insights to inform person-centred and compassionate care. Nevertheless, it must also be recognised that this is a small sample of patients and families rather than a representative of the diverse population we serve. This work needs to evolve through continued service evaluation and improved strategies for capturing and understanding the experience of underserved populations (NHS England, No Date).

### Next steps

Service evaluation engagement events have been pivotal in informing service improvement and engagement strategies within critical care. The feedback, once checked for accuracy with the participants, was shared with the entire critical care team. In addition to these events, staff listening events were conducted to triangulate the experiences of patients, families, and staff.

As a department, multiple members of the multidisciplinary team have received experience-based co-design training and are using a proactive approach to patient and public engagement and involvement, recruiting participants once they have been discharged to other wards in the hospital.

The Critical Care Patient and Family team, in collaboration with the Critical Care Data Clinic, are analysing health inclusion data from electronic health records to gain a deeper understanding of the diverse population of patients and families we serve. This will enable us to proactively collect experiential data that is more representative of the population we serve.

## Data Availability

All data produced in the present study are available upon reasonable request to the authors

## Acknowledgements

The authors would like to acknowledge Siri Steinmo, Elizabeth Taylor, Jeyapragash Jeyapala, Kathleen Thomas, Jacob Levi, Philippa Guppy, Victoria Dunne, Teona Serafimova, Talhah Atcha, Anna Welch, and Emily Walker for their work evaluating the FLT in 2021 that led to the development of the Critical Care Inpatient Journey (figure 1.). Also, Meena Patel, Lisa Anderton and Rossana Fazzina for their assistance in organising the events. Special thanks to all patients and family participants.

